# Effect of COVID-19 on Critical ICU Capacity in US Acute Care Hospitals

**DOI:** 10.1101/2020.12.16.20248366

**Authors:** Thomas C. Tsai, Benjamin H. Jacobson, Ashish K. Jha

**Affiliations:** Department of Health Policy and Management, Harvard Chan School of Public Health, Boston, MA; Department of Surgery, Brigham and Women’s Hospital, Boston, MA; Brown University School of Public Health, Providence, RI

**Author notes:** Corresponding Author: Thomas C. Tsai, MD, MPH, Brigham and Women’s Faulkner Hospital, 1153 Centre Street, Boston, MA 02130.

## Abstract

**Importance:** The current wave of COVID-19 infections has led to media reports of ICUs across the country reaching critical capacity. But the degree to which this has happened and community and institutional characteristics of hospitals where capacity limits have been reached is largely unknown.

**Objective:** To determine changes in intensive care capacity in US acute care hospitals between September and early December, 2020 and to identify whether hospitals serving more vulnerable populations were more likely to exceed critical-levels of ICU occupancy.

**Design, Setting, and Participants:** Retrospective observational cohort of US acute care hospitals reporting to the US Department of Health and Human Services (HHS) from September 4, 2020 to December 3, 2020. Hospitals in this cohort were compared to all US acute care hospitals. Multivariate logistic regression was used to assess the relationship between community socioeconomic factors and hospital-structural features with a hospital reaching critical ICU capacity.

**Exposure:** Community-level socioeconomic status and hospital-structural features

**Main Outcomes and Measures:** Our primary outcome was reaching critical ICU capacity (>90%) for at least two weeks since September 4. Secondary outcomes included the weekly capacity and occupancy tabulated by state and by hospital referral region.

**Results:** 1,791 hospitals had unsuppressed ICU capacity data in the HHS Protect dataset, with 45% of hospitals reaching critical ICU capacity for at least two weeks during the study period. Hospitals in the South (OR = 2.79, p<0.001), Midwest (OR = 1.76, p=0.01) and West (OR = 1.85, p<0.01) were more likely to reach critical capacity than those in the Northeast. For-profit hospitals (OR = 2.15, p<0.001), rural hospitals (OR = 1.40, p<0.05) and hospitals in areas of high uninsurance (OR = 1.94, p<0.001) were more likely to reach critical ICU capacity, while hospitals with more intensivists (OR = 0.92, p=0.044 and higher nurse-bed ratios (OR = 0.95, p=0.013) were less likely to reach critical capacity.

**Conclusions and Relevance:** Nearly half of U.S. hospitals reporting data to HHS Protect have reached critical capacity at some point since September. Those that are better resourced with staff were less likely to do so while for for-profit hospitals and those in poorer communities were more likely to reach capacity. Continued non-pharmacologic interventions are clearly needed to spread of the disease to ensure ICUs remain open for all patients needing critical care.

**Key Points:** *Question:* **With an increasing number of SARS-CoV2 infections**, how has the burden on ICU capacity changed over the past three months and what community and institutional factors are associated with hospitals reaching critical capacity?

*Finding:* 45% of US acute care hospitals have reached critical ICU capacity at some point over the past three months. Hospital located in areas with fewer insured people were more likely to reach critical ICU capacity. At an institutional level, for-profit hospitals, rural hospitals, and those that have less baseline staffing of intensivists and nurses were more likely to reach critical ICU capacity.

*Meaning:* The COVID-19 pandemic appears to be disproportionately straining ICUs with fewer resources and staff, setting up a substantial risk to widen disparities in access to care for already underserved populations.

---

As SARS-CoV2 continues to infect more than 200,000 Americans daily, the clinical impact of all of those infections are starting to be felt by acute care hospitals. Media stories of hospital overcrowding and struggling to manage the surge of stories have become legion. The ability acute care hospitals to deliver timely, effective care is fundamental not just for patients with the novel 2019 coronavirus disease (COVID-19) but also for everyone else who relies on hospital care.

Tracking real-time data on hospital ICU care is critically important but has largely been unavailable at the individual hospital level, hampering our ability to fully understand how effectively our hospitals are able to manage the influx of patients. Initial voluntary data collection by the Centers for Disease Control and Prevention National Healthcare Safety Network had both low reporting rates and was not easily inaccessible. For the first time since the start of the pandemic, detailed hospital-level data on hospital capacity is available with the public release of this HHS Protect data on December 7, 2020. This opens up the possibility that we can now understand how widespread pressures on hospital ICUs are, and the factors that may be driving why some hospitals are more stretched than others. There is reason to believe it will vary. It may be that some types of hospitals may be less likely to postpone elective surgeries, thus risking higher levels of ICU utilization. Other institutions may serve a primarily vulnerable population who are more likely to get infected and sick, and therefore those institutions may be more likely to see ICU capacity strains. Empirical data on those questions would be immensely helpful.

Using newly released hospital-level ICU capacity data, we sought to answer three pressing policy questions. First, what has been the trend in ICU capacity over the current phase of the pandemic and how has it varied across US hospital markets. Second, given known disparities in medical access, what are the community-level features associated with hospitals with critical ICU capacity gaps? Lastly, what are the structural and staffing features of hospitals with critical ICU capacity gaps. Specifically, we hypothesized that rural hospitals, due to low bed supply, and for-profit hospitals, due to financial incentives, may be more likely to be susceptible to critical ICU capacity during the current phase of the COVID-19 pandemic.

## Methods

### Data

We merged together several data sets in order to assess the community and hospital-level factors associated with critical ICU capacity. The HHS Protect Public Data Hub consolidates multiple data sources from HHS and CDC to create a dashboard of hospital capacity in the US in response to COVID-19. Additionally, HHS requires all hospitals licensed to provide 24-hour care to report data to the HHS Protect Effort. Initial dashboards were only at the state level, but a hospital-level public dataset was released on December 7, 2020. The HHS Protect hospitalization data file was merged with the 2018 American Hospital Association (AHA) Annual Survey using hospital CMS certification numbers (CCN) to obtain hospital-level structural features. The Area Health Resource File landscape file was then merged to the analytic dataset using county FIPS codes to obtain county-level socioeconomic variables. Hospitals in the top decile of disproportionate share index were defined as safety net hospitals. Data points smaller than four were suppressed by HHS and these hospital-week observations were excluded from analysis. 117 of the 4,812 hospitals in the dataset did not have observations for all 13 weeks since September 1, 2020. 33 of these hospitals were missing more than four weeks of data and were excluded.

### Variables

Our main dependent variable is a hospital reaching critical ICU capacity, which was defined as a hospital’s ICU occupancy exceeding 90% capacity for at least two weeks since September 1. We chose overall ICU capacity for both COVID and non-COVID-related admissions as our main dependent variable as it reflects the effect of the burden of COVID-19 on the overall ability of acute hospitals to deliver timely and effective care for the sickest patients. Hospital occupancy for each week was determined by dividing the number of occupied ICU beds by the number of staffed ICU beds.

We had two sets of independent variables—community and hospital-level factors. Our main predictor was the hospital-level factors of for-profit status and rural status. Community-level variables were geographic region, median age in county, percent uninsured in county, proportion Black in county, and percent Hispanic in county. Age, uninsured, Black, and Hispanic were converted into quartiles across the data. Hospital-level variables explored for association with reaching critical ICU capacity were hospital size (less than 100 beds is small, 100-399 beds is medium, 400 or more beds is large), teaching status (non-teaching, major teaching, or minor teaching), rurality, profit status, safety net status, nurse-bed ratio, number of intensivists, and number of operating rooms. Rurality was defined based on the CMS definition using core-based statistical area (CBSA). Safety-net was defined as the top decile of the disproportionate share (DSH) index which was obtained from the 2017 CMS Healthcare Cost Report Information System (HCRIS).

### Analysis

We first compared the characteristics of the HHS Protect sample to the overall sample of acute care hospitals in the AHA Annual Survey. Our main analyses included only the set of hospitals that had available and unsuppressed ICU data. To determine a trend of critical ICU capacity, ICU occupancy and capacity were aggregated across all hospitals and stratified by week and by region to determine national and regional trends in ICU occupancy. Regions were divided using US Census Bureau categorizations as follows: Northeast (CT, MA, ME, NH, RI, VT, NJ, NY, PA), Midwest (OH, IN, IL, MI, WI, MN, IA, MO, KS, NE, SD, ND), South (KY, TN, MS, AL, AR, LA, OK, TX, DE, DC, FL, GA, MD,NC, SC, VA, WV) and West (MT, ID, WY, CO, NM, AZ, UT, NV, WA, OR, CA, AK, HI). Occupancy in the most recent week of data was aggregated by Hospital Referral Region (HRR) and displayed in a heatmap to determine geographic variation in occupancy. Occupancy was also tabulated at the state level.

We next assessed the relationship between community-level factors and a hospital reaching critical ICU capacity. Mean values of hospital- and community-level factors were calculated independently for ICUs that did and did not exceed 90% capacity for at least two weeks since September 1. Community-level factors were assessed for a relationship with reaching critical ICU capacity using bivariate logistic regressions. A multivariate logistic regression model at the hospital-level was then created to assess the relationship between community-level factors and a hospital reaching critical ICU capacity.

In order to assess the main relationship between hospital-level factors and a hospital reaching critical ICU capacity, we first compared bivariate relationships. We then created a multivariate logistic regression model to assess the relationship between hospital-level factors and a hospital reaching critical ICU capacity. All analyses were performed using R Version 3.6.2. This study was exempt by the institutional review board of the Harvard T. H. Chan School of Public Health

## Results

A total of 4,327 hospitals were included in the HHS Protect dataset, of which 1,791 had unsuppressed ICU capacity data. Hospitals with less than 4 ICU beds were suppressed, so the resulting sample substantial underrepresented small and rural hospitals (**Supplementary Table 1**). Critical access hospitals were especially affected by data suppression, with only 4 of 1,282 critical access hospitals having available ICU capacity data.

ICU occupancy has increased across the US, though the upward trend has been most substantial in the South and Midwest (**Figure 1**). In the South, ICU capacity increased by about 10% (30,523 to 33,142) while occupancy grew by 16% (22,872 to 26,624) resulting in overall occupancy growing from 74.9% to 80.3%. In the Midwest, capacity remained stagnant while occupancy surged (11,285 to 14,210), bringing overall occupancy from 62.0% to 74.8%. In line with this increasing overall occupancy, the proportion of hospitals in the Midwest with ICUs at critical capacity – defined as having more than 90% of beds occupied – nearly tripled from 10% in September to 28% by the end of November. The South (26% to 42%), West (22% to 30%) and Northeast (8% to 18%) also saw the proportion of hospitals at critical ICU capacity increase from September to early December. Since mid-October, much of this increase has been driven by COVID cases, with COVID-occupied ICU beds nationwide doubling from 9,507 to over 21,203 in just five weeks. Total ICU occupancy increased by only 4,672 beds over the same five weeks, suggesting that non-COVID ICU cases decreased.

**Figure 1.**
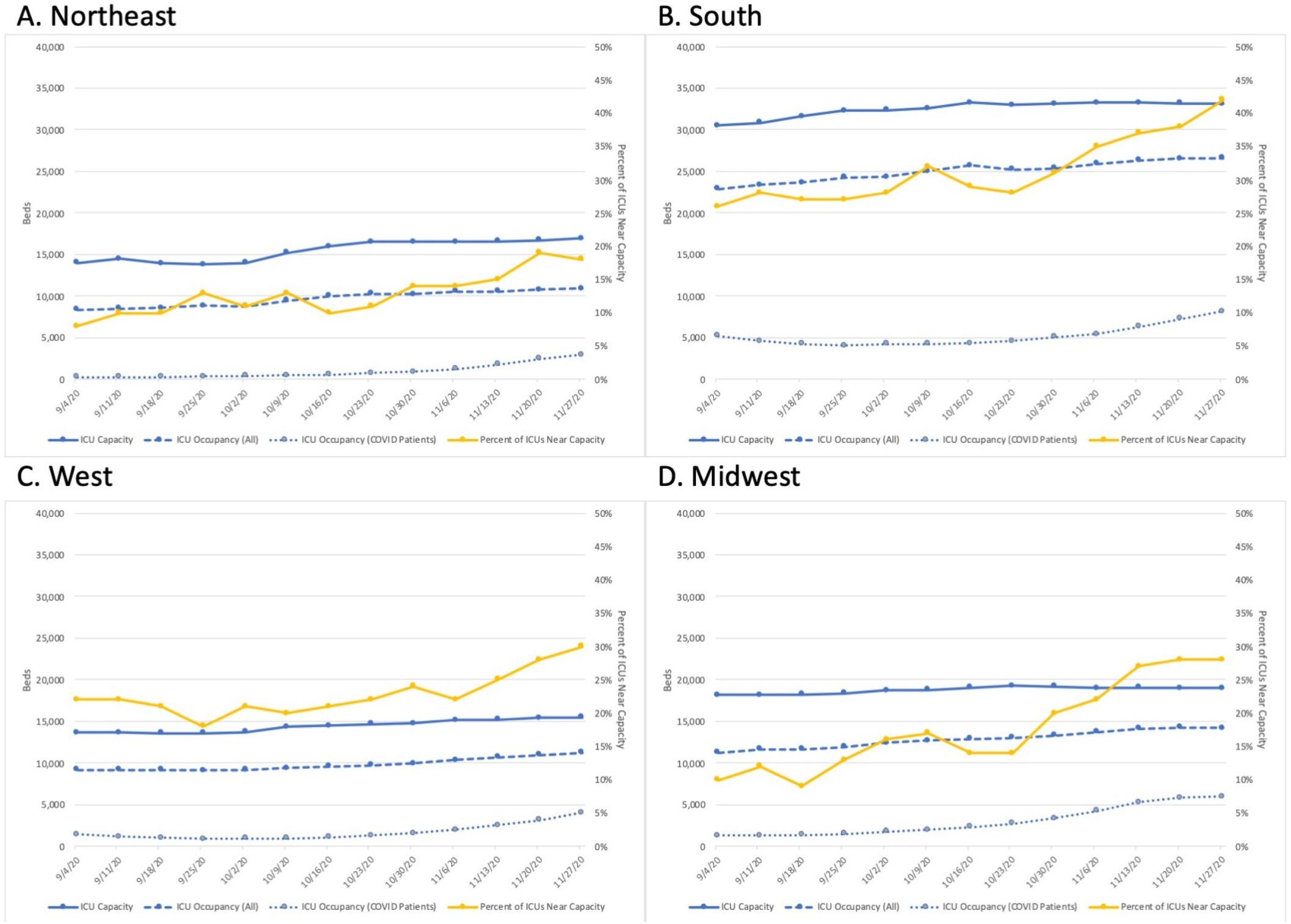
Trend in Regional US ICU Occupancy and Capacity over the Third Phase of the COVID-19 Pandemic, September to December 2020

The distribution of US acute care hospitals reaching critical ICU capacity during the COVID-19 pandemic shows substantial geographic variation (**Figure 2**). In the most recent week of available data (November 27 to December 4), hospital referral regions (HRRs) in the South and Midwest appeared most overburdened, while the Northeast and West Coast showed lower ICU occupancy. The HRRs with the highest occupancies were Albuquerque, NM, Ogden, UT, Oxford, MS, St. Cloud, MN, and St. Joseph, MI, all of which saw ICU occupancy at or over 100% of their reported ICU capacity (**Supplemental Table 2**). In contrast, the HRRs with the lowest occupancy rates were Bronx, NY, Rochester, NY, Greeley, CO, Buffalo, NY, and Springfield, MA, all of which had fewer than 50% of reported ICU capacity occupied. Similar results were observed when tabulating ICU occupancy by state, with no state dipping below 50% occupancy. (**Supplemental Table 3**).

**Figure 2.**
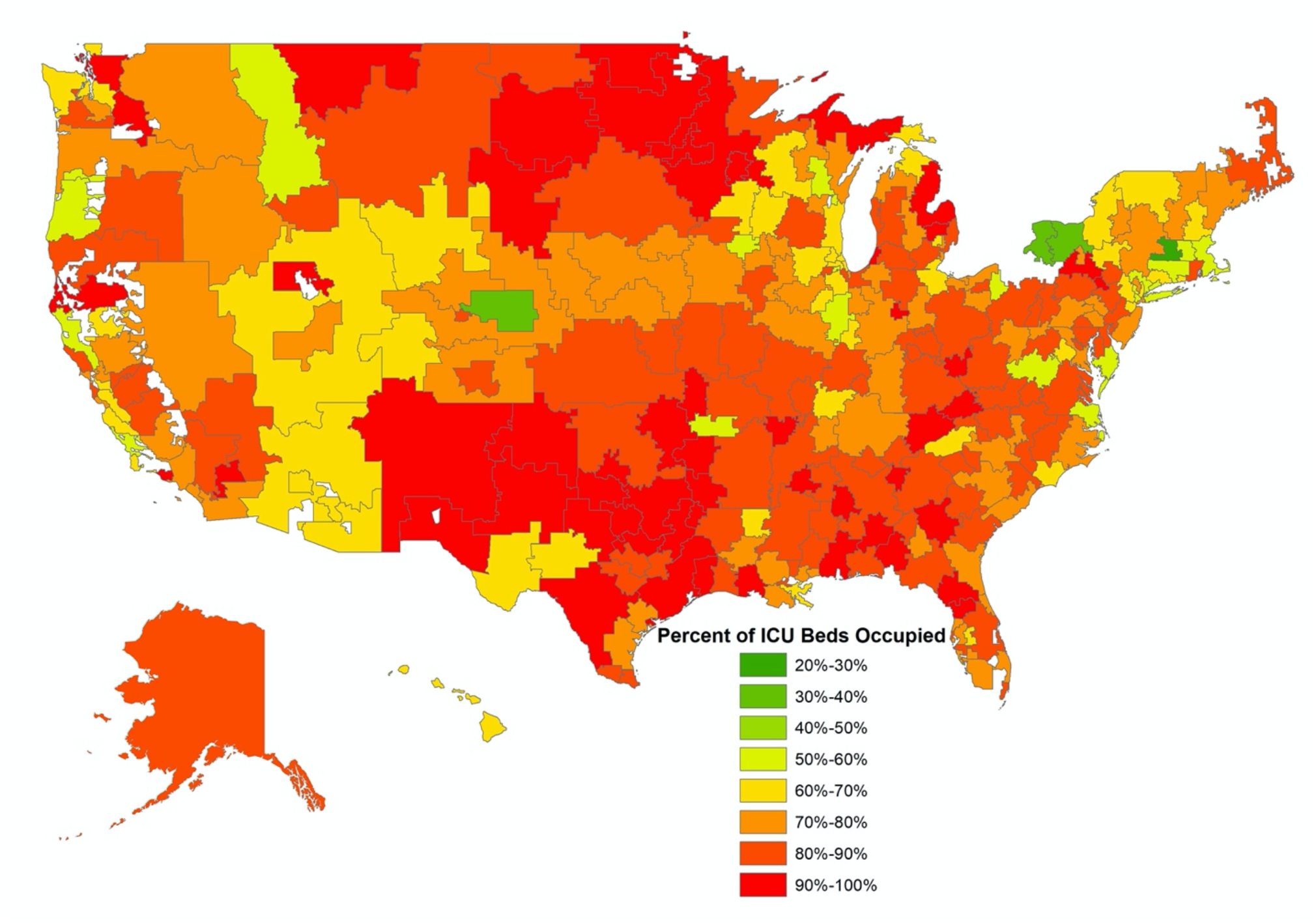
HRR-Level Geographic Variation in ICU Occupancy for the Week of November 27 HRRs in which all hospitals had data suppressed do not appear on this map.

Of the 1,791 hospitals with ICU occupancy data, 45% (805) reached critical ICU capacity for at least two weeks since September, and these hospitals differed from those that did not reach critical capacity on a number of county- and hospital-level factors (**Supplemental Table 4**). Regional effects strongly influenced the likelihood of hospitals reaching critical capacity, with hospitals in the South having almost 3 times the odds (OR = 2.79, p < 0.001) of hitting critical capacity as those in the Northeast (**Table 1**). Hospitals in the Midwest and West also had increased odds (OR = 1.76, 1.85, p = 0.002, 0.004, respectively) of reaching critical capacity relative to those in the Northeast. Hospitals in counties with a high percentage of uninsured residents had higher odds of hitting critical capacity, with those in the fourth quartile of uninsurance having nearly double the odds of reaching critical capacity as those in the first quartile (OR = 1.94, p = 0.001). Higher median age in a county appeared to reduce odds of reaching critical capacity in a bivariate model, though in a multivariate model with p value corrections for multiple hypothesis testing, this relationship no longer appeared significant. The proportion of a county that identified as Black or Hispanic did not appear to impact the likelihood of hospitals within that county exceeding capacity.

**Table 1.**
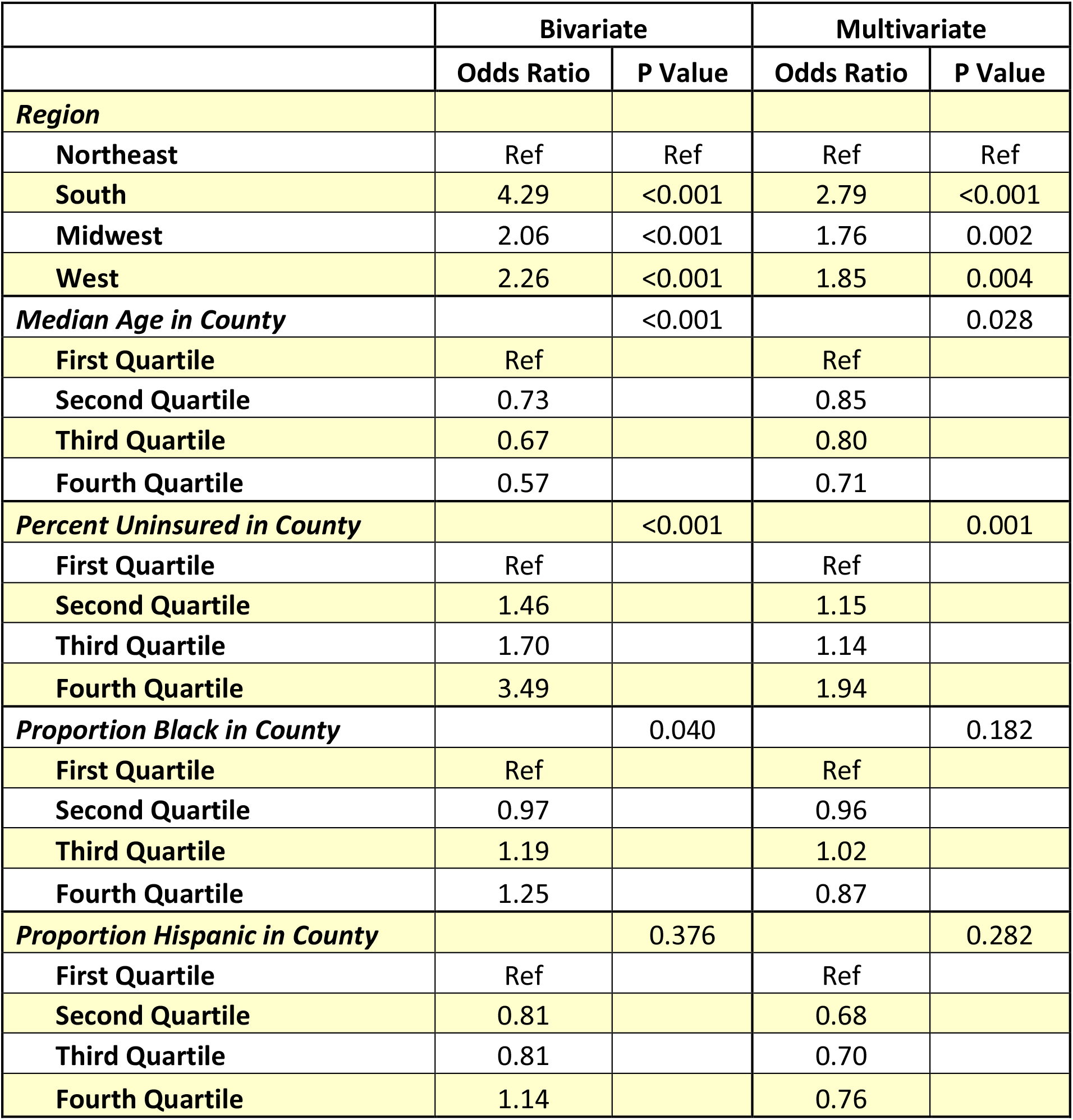
Association between Community-Level Factors and Odds of Reaching Critical ICU Capacity

|  | **Bivariate** |  | **Multivariate** |  |
| --- | --- | --- | --- | --- |
|  | **Odds Ratio** | **P Value** | **Odds Ratio** | **P Value** |
| ***Region*** |  |  |  |  |
| Northeast | Ref | Ref | Ref | Ref |
| South | 4.29 | <0.001 | 2.79 | <0.001 |
| Midwest | 2.06 | <0.001 | 1.76 | 0.002 |
| West | 2.26 | <0.001 | 1.85 | 0.004 |
| ***Median Age in County*** |  | <0.001 |  | 0.028 |
| First Quartile | Ref |  | Ref |  |
| Second Quartile | 0.73 |  | 0.85 |  |
| Third Quartile | 0.67 |  | 0.80 |  |
| Fourth Quartile | 0.57 |  | 0.71 |  |
| ***Percent Uninsured in County*** |  | <0.001 |  | 0.001 |
| First Quartile | Ref |  | Ref |  |
| Second Quartile | 1.46 |  | 1.15 |  |
| Third Quartile | 1.70 |  | 1.14 |  |
| Fourth Quartile | 3.49 |  | 1.94 |  |
| ***Proportion Black in County*** |  | 0.040 |  | 0.182 |
| First Quartile | Ref |  | Ref |  |
| Second Quartile | 0.97 |  | 0.96 |  |
| Third Quartile | 1.19 |  | 1.02 |  |
| Fourth Quartile | 1.25 |  | 0.87 |  |
| ***Proportion Hispanic in County*** |  | 0.376 |  | 0.282 |
| First Quartile | Ref |  | Ref |  |
| Second Quartile | 0.81 |  | 0.68 |  |
| Third Quartile | 0.81 |  | 0.70 |  |
| Fourth Quartile | 1.14 |  | 0.76 |  |

In addition to county characteristics, hospital characteristics were assessed for a relationship with odds of reaching critical capacity (**Table 2**). For-profit hospitals had considerably higher odds of hitting critical capacity than non-profit hospitals (OR = 2.15, p < 0.001). Rural hospitals also appeared to have higher odds of reaching critical capacity (OR = 1.46, p = 0.040). Hospitals with more intensivists (8% lower odds for each additional 10 intensivists) and higher nurse-bed ratios (5% lower odds for each one unit increase in nurse-bed ratio) had lower odds of reaching critical capacity than those with lesser staffing (OR= 0.92, 0.95, P=0.044, 0.013, respectively). Safety-net status, teaching status, the number of operating rooms, and the size of hospitals all showed no significant relationship with the odds of hitting critical ICU capacity.

**Table 2.** Association between Hospital-Level Factors and Odds of Reaching Critical ICU Capacity

|  | **Bivariate** |  | **Multivariate** |  |
| --- | --- | --- | --- | --- |
|  | **Odds Ratio** | **P Value** | **Odds Ratio** | **P Value** |
| ***Size*** |  |  |  |  |
| Small | Ref | Ref | Ref | Ref |
| Medium | 0.94 | 0.561 | 1.09 | 0.540 |
| Large | 1.07 | 0.773 | 1.37 | 0.369 |
| ***Teaching Status*** |  |  |  |  |
| Non-Teaching | Ref | Ref | Ref | Ref |
| Major Teaching | 0.91 | 0.536 | 1.07 | 0.765 |
| Minor Teaching | 0.97 | 0.790 | 0.95 | 0.681 |
| Rural Hospital | 1.47 | 0.010 | 1.46 | 0.040 |
| For Profit Hospitals | 1.80 | <0.001 | 2.15 | <0.001 |
| Safety Net Hospital | 0.92 | 0.568 | 0.93 | 0.722 |
| Nurse-Bed Ratio | 0.94 | <0.001 | 0.95 | 0.013 |
| Intensivists (per 10) | 0.92 | 0.012 | 0.92 | 0.044 |
| Operating Rooms | 1.00 | 0.858 | 1.00 | 0.554 |

## Discussion

In this analysis of newly available official national acute-care hospital ICU occupancy data, we found that almost half of hospitals reached critical ICU capacity, exceeding 90% occupancy for at least two weeks during this phase of the COVID-19 pandemic. Hospitals in the South and Midwest have grown more overburdened than the rest of the country, though rising occupancy has impacted every region. Hospitals in more vulnerable communities, as measured by rates of uninsurance, were far more likely to reach ICU capacity as were for-profit hospitals and those that were located in rural region. Finally, we found that well-staffed hospitals with more intensivists and more nurses were less likely to approach critical ICU capacity. Taken together, these results highlight the growing crisis of overburdened ICUs across the country and point to the fact that staffing and resources are likely central factors in hospitals facing the greatest threats.

We found that the surge of COVID-19 cases in the ICU far outpaced the overall growth of ICU occupancy, suggesting that fewer non-COVID patients were being admitted to the ICU during this time. This is consistent with stories of more restrictive criteria for hospitalizations and ICU care in the face of shrinking available capacity. Such changes driven by rationing of care due to rising severe COVID-19 cases essentially lead to “crowding out” non-COVID-related but necessary medical care. Recent findings show that a substantial proportion of excess mortality over the past months can be attributed to non-COVID causes such as heart disease,^8^ and the reduction of ICU admittances for non-COVID illnesses may play a potent role in this excess mortality. If this change in ICU admittance criteria were indeed leading to worsening illness and excess mortality, it would provide another mechanism by which COVID surges can harm public health, by making care for other conditions harder to obtain.

While ICU capacity has moderately expanded over the past few months, this growth has failed to keep pace with rising caseloads. Our study highlights the importance of slowing community-level transmission to prevent hospitals from being overwhelmed. In the early months of the pandemic, ICU capacity was greatly limited by a shortage of ventilators and PPE,^9^ which hampered any attempts to convert general hospital beds into ICU beds. A massive manufacturing effort has since grown the ventilator supply far beyond the needed capacity,^10^ and yet the American ICU capacity has still proved largely inelastic. One potential cause for this inelasticity is a shortage in the critical care workforce. A number of studies have documented worse outcomes in ICUs with lower nurse-patient ratios,^11,12^ and simply operating the numerous ventilators required by a surge of COVID-related ARDS necessitates a sufficient quantity of intensivists. We find lower odds of reaching critical capacity in hospitals with greater numbers of intensivists and more nurses. While finding and adding ventilators and other equipment can help with expanding ICU capacity, it becomes increasingly difficult to obtain more trained staff.

One way hospitals have been able to add capacity is by cancelling so-called elective procedures and maximizing beds allocated for COVID-19 patients. Cancelling such procedures can prove very costly from a financial perspective,^13^ though the benefits from a capacity standpoint are such that CMS recommended and most states mandated such closures in March and April.^14^ There have been no federal recommendations and few state mandates during the current surge, leaving decisions about cancelling procedures to individual hospitals. Our finding that for-profit hospitals had more than double the odds of reaching critical capacity as non-profit hospitals may be a results of these for-profit hospitals possibly being less willing to forego elective procedures and may therefore have failed to create sufficient capacity for the surge of severe COVID-19 cases. For-profit hospitals may also be less willing to transfer patients to other hospitals, balancing ICU burdens but giving up sources of future revenue.

ICU capacity has grown on average over the past 20 years but much of this growth has been concentrated in urban areas, with rural hospitals seeing little change in capacity.^15^ Indeed the acceleration of rural hospital closures in recent years means rural areas may have entered the pandemic with less ICU capacity than at any point in the previous decade.^16^ With hospital closures forcing remaining hospitals to care for larger populations, our finding of rural hospitals with higher odds of reaching critical ICU capacity highlights the need to support rural hospitals. Similar disparities in access to ICU capacity exist in low-income areas with high rates of uninsurance^17^. Lack of insurance may be causing greater surges in these communities either directly, because people delay seeking care, or as a proxy for broader social challenges that lead to more multi-generational homes, jobs that are hard to do socially-distanced, and as a result, more spread of the disease in those communities. These findings demonstrate the particular vulnerability of these communities to the COVID-19 pandemic. Current efforts and future pandemic preparedness and response should direct resources to bolster access to acute hospital care for patients residing in rural counties and counties with higher rates of poverty.

## Limitations

The HHS Protect hospitalization dataset suppresses small capacity and occupancy numbers, which leads to a substantial underrepresentation of rural and critical access hospitals. Our findings may underestimate the impact of rurality on hospital occupancy, and we were unable to analyze whether critical access hospitals had higher odds of reaching critical ICU capacity. The suppression of occupancy data on critical access hospitals in the HHS Protect dataset is puzzling—while the need to protect patient privacy is important in hospitals with low volume of discharges, highlighting the capacity gaps in critical access hospitals should be a priority for US policymakers. Data on these hospitals would be valuable for future analysis to understand the risks faced by the most vulnerable hospitals. However, these hospitals represent a small fraction of the ICU beds in the United States and our findings still hold for the majority of ICUs caring for COVID-19 patients. There is little data on ICU occupancy and capacity in the early months of the pandemic, and this study is limited to the third wave of the pandemic starting in September 2020. Given how much the challenges faced by hospitals and ICUs have evolved over the pandemic, however, it may be more informative to focus on the current surge of cases and this current analysis may be useful to policymakers and hospital administrators.

## Conclusion

In conclusion, we find a substantial shortage of ICU capacity in the US, with nearly 1 in 2 hospitals in the US exceeding 90% capacity for at least two weeks during the current wave of the COVID-19 pandemic. This growing crisis of overburdened ICUs has been driven by surging COVID-19 cases and has led to a reduction in non-COVID ICU care. Our finding of an association between higher staffing and lower odds of reaching critical capacity supports the importance of the critical care workforce in ensuring adequate capacity to care for all COVID-19 patients. Hospitals in rural and high-uninsured communities had greater risk of reaching critical capacity, demonstrating an exacerbation of healthcare disparities by the COVID-19 pandemic. As COVID-19 caseloads continue to grow, policymakers should bolster support for hospitals serving vulnerable communities.

## Data Availability

HHS Protect Data is available online

https://protect-public.hhs.gov/pages/covid19-module

---

**Supplemental Table 1.** Comparison Between All ACA Hospitals and Hospitals with Unsuppressed Data

|  | **AHA Hospitals** | **Hospitals with ICU**<br>**Data** |
| --- | --- | --- |
| **Total Hospitals** | 4327 | 1791 |
| ***Size of Hospital*** |  |  |
| **Small** | 3171 (0.73) | 792 (0.44) |
| **Medium** | 1059 (0.24) | 910 (0.51) |
| **Large** | 97 (0.02) | 89 (0.05) |
| ***Teaching Status (Proportion)*** |  |  |
| **Non-Teaching** | 2992 (0.69) | 855 (0.48) |
| **Minor Teaching** | 1097 (0.06) | 725 (0.40) |
| **Major Teaching** | 238 (0.25) | 211 (0.12) |
| ***Region (Proportion)*** |  |  |
| **Northeast** | 542 (0.13) | 295 (0.16) |
| **Midwest** | 1316 (0.30) | 398 (0.22) |
| **South** | 1617 (0.37) | 746 (0.42) |
| **West** | 852 (0.20) | 352 (0.20) |
| **Rural Hospitals (Proportion)** | 1749 (0.40) | 206 (0.12) |
| **Critical Access Hospitals (Proportion)** | 1282 (0.30) | 4 (0.00) |

**Supplemental Table 2.** Ordering of HRRs by ICU Occupancy for the Week of November 27

| **HRR** | **Staffed**<br>**ICU**<br>**Beds \*** | **Occupied**<br>**ICU Beds \*** | **Proportion of**<br>**Hospitals with**<br>**ICUs at Critical**<br>**Capacity** | **ICU**<br>**Occupancy** |
| --- | --- | --- | --- | --- |
| Ogden, UT | 58.4 | 60.2 | 0.67 | 1.03 |
| Oxford, MS | 14 | 14.3 | 1.00 | 1.02 |
| St. Cloud, MN | 48.4 | 48.4 | 1.00 | 1.00 |
| St. Joseph, MI | 20.4 | 20.4 | 1.00 | 1.00 |
| Wichita Falls, TX | 36.1 | 35.9 | 1.00 | 0.99 |
| Rapid City, SD | 30.9 | 30.7 | 1.00 | 0.99 |
| Victoria, TX | 12.1 | 11.9 | 1.00 | 0.98 |
| Beaumont, TX | 95.8 | 94 | 1.00 | 0.98 |
| ... | ... | ... | ... | ... |
| Salem, OR | 66 | 33.4 | 0.00 | 0.51 |
| Appleton, WI | 31.3 | 15.7 | 0.00 | 0.50 |
| San Luis Obispo, CA | 37.3 | 18.7 | 0.00 | 0.50 |
| Bronx, NY | 595.7 | 247.5 | 0.00 | 0.42 |
| Rochester, NY | 617.3 | 217.9 | 0.00 | 0.35 |
| Greeley, CO | 124 | 42.2 | 0.00 | 0.34 |
| Buffalo, NY | 730.6 | 242.7 | 0.00 | 0.33 |
| Springfield, MA | 235.6 | 66 | 0.00 | 0.28 |

\* Weekly Average

**Supplemental Table 3.**
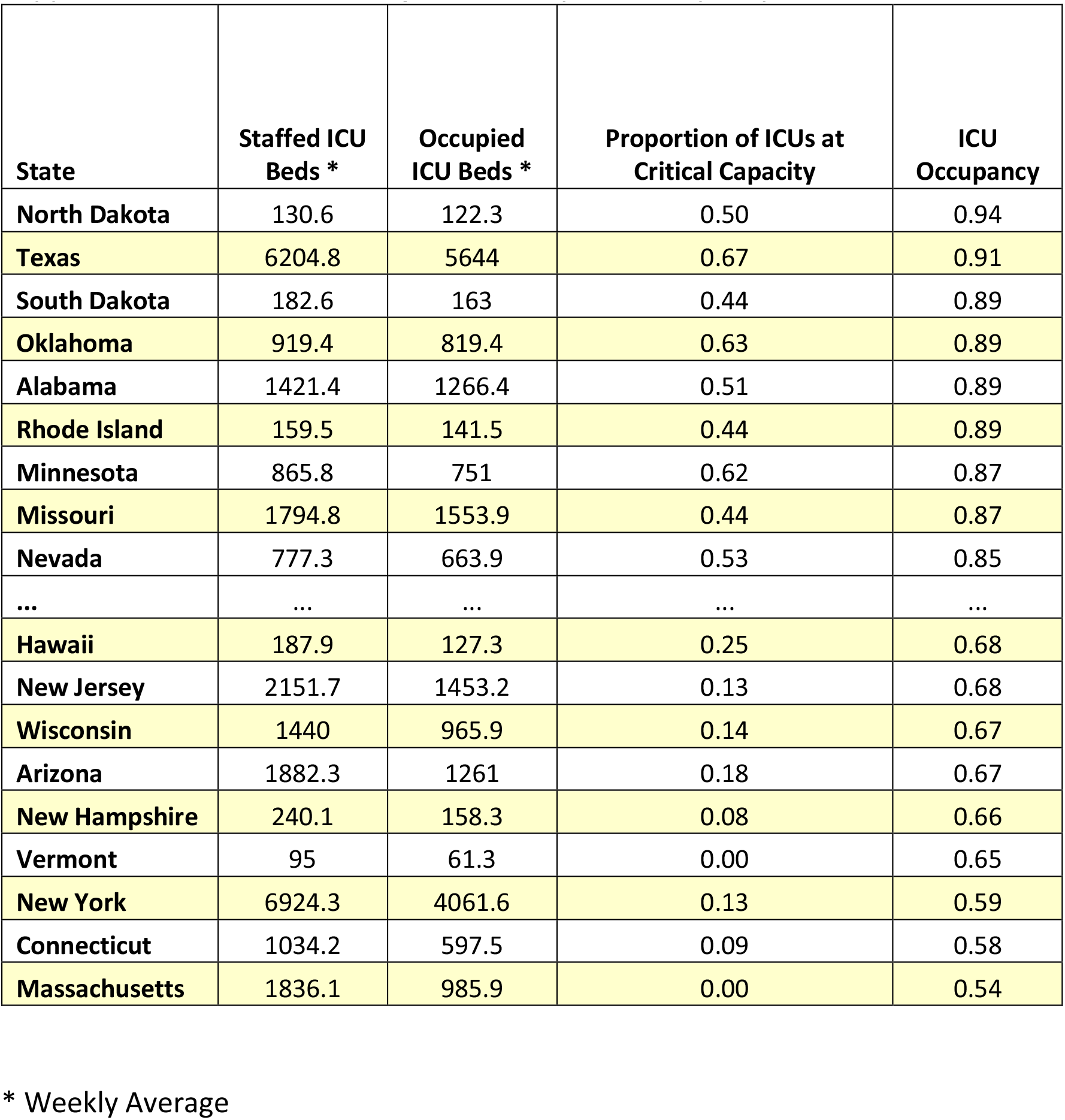
Ordering of States by ICU Occupancy for the Week of November 27

| State | Staffed ICU Beds * | Occupied ICU Beds * | Proportion of ICUs at Critical Capacity | ICU Occupancy |
| --- | --- | --- | --- | --- |
| North Dakota | 130.6 | 122.3 | 0.50 | 0.94 |
| Texas | 6204.8 | 5644 | 0.67 | 0.91 |
| South Dakota | 182.6 | 163 | 0.44 | 0.89 |
| Oklahoma | 919.4 | 819.4 | 0.63 | 0.89 |
| Alabama | 1421.4 | 1266.4 | 0.51 | 0.89 |
| Rhode Island | 159.5 | 141.5 | 0.44 | 0.89 |
| Minnesota | 865.8 | 751 | 0.62 | 0.87 |
| Missouri | 1794.8 | 1553.9 | 0.44 | 0.87 |
| Nevada | 777.3 | 663.9 | 0.53 | 0.85 |
| ... | ... | ... | ... | ... |
| Hawaii | 187.9 | 127.3 | 0.25 | 0.68 |
| New Jersey | 2151.7 | 1453.2 | 0.13 | 0.68 |
| Wisconsin | 1440 | 965.9 | 0.14 | 0.67 |
| Arizona | 1882.3 | 1261 | 0.18 | 0.67 |
| New Hampshire | 240.1 | 158.3 | 0.08 | 0.66 |
| Vermont | 95 | 61.3 | 0.00 | 0.65 |
| New York | 6924.3 | 4061.6 | 0.13 | 0.59 |
| Connecticut | 1034.2 | 597.5 | 0.09 | 0.58 |
| Massachusetts | 1836.1 | 985.9 | 0.00 | 0.54 |

\* Weekly Average

**Supplemental Table 4.** Summary Hospital- and County-Level Characteristics for Hospitals that Did and Did Not Reach Critical ICU Capacity

|  | ICU at<br>Critical<br>Capacity | ICU Not at<br>Critical<br>Capacity |
| --- | --- | --- |
| **Total Hospitals** | 805 (0.45) | 986 (0.55) |
| ***Region*** |  |  |
| **Northeast** | 71 (0.09) | 224 (0.23) |
| **Midwest** | 157 (0.2) | 241 (0.24) |
| **South** | 430 (0.53) | 316 (0.32) |
| **West** | 147 (0.18) | 205 (0.21) |
| **Average Median Age in County** | 36.7 | 37.5 |
| **Average Percent Uninsured in County** | 11.6 | 9.3 |
| **Average Percent Black in County** | 11 | 10.2 |
| **Average Percent Hispanic in County** | 7.1 | 6.2 |
| ***Size of Hospital*** |  |  |
| **Small** | 361 (0.45) | 431 (0.44) |
| **Medium** | 402 (0.5) | 508 (0.52) |
| **Large** | 42 (0.05) | 47 (0.05) |
| ***Teaching Status*** |  |  |
| **Non-Teaching** | 389 (0.48) | 466 (0.47) |
| **Minor Teaching** | 325 (0.4) | 400 (0.41) |
| **Major Teaching** | 91 (0.11) | 120 (0.12) |
| **Rural Hospital** | 110 (0.14) | 96 (0.1) |
| **For Profit Hospitals (Proportion)** | 195 (0.24) | 149 (0.15) |
| **Safety Net Hospitals** | 86 (0.11) | 109 (0.11) |
| **Nurse-Bed Ratio** | 8.9 | 9.5 |
| **Average Number of Intensivists** | 4.3 | 6.2 |
| **Average Number of ORs** | 17.3 | 17.4 |

